# Executive Function and Adherence in Children and Adolescents Living with HIV: Evidence from the HIV-associated Neurocognitive Disorders in Zambia (HANDZ) Study

**DOI:** 10.1101/2024.09.17.24313838

**Authors:** Sylvia Mwanza-Kabaghe, Kristen Sportiello, Mina Shah, Heather R. Adams, Esau G. Mbewe, Pelekelo P. Kabundula, Colleen Schneider, Milimo Mweemba, Gretchen L. Birbeck, David R. Bearden

## Abstract

**Introduction:** Executive function (EF) may be impaired in people with human immunodeficiency virus (HIV) infection, and poor EF may affect medication adherence. However, there is little data on EF in children with HIV in sub-Saharan Africa.

**Methods:** 208 children/adolescents with perinatally acquired HIV and 208 HIV-exposed uninfected controls were recruited in Zambia for this prospective cohort study. EF was measured using performance-based, self-report, and parental report measures. Adherence over one year of follow-up was assessed through questionnaires and viral load measurement.

**Results:** Children with HIV performed significantly worse on all three measures of EF. Lower parental rating of EF was associated with poorer antiretroviral therapy adherence (OR: 1.5, 95% CI = 1.02 – 2.2, p = 0.04).

**Conclusion:** Children with HIV have EF impairments which may lead to consequences like poor medication adherence and treatment failure. Interventions to improve EF or compensate for impaired EF may be necessary in this population.

## Introduction

Executive function (EF) includes a suite of cognitive skills including attention, planning, and behaviour control which are critical for educational and professional success, as well as facilitating activities such as medication adherence.^1–4^ A substantial body of research has demonstrated impairment in executive function in people living with HIV,^5–32^ although the precise mechanism for this impairment and the extent to which it affects function is unknown. With the widespread deployment of antiretroviral therapy (ART), children and adolescents with perinatally acquired HIV are now surviving into adulthood.^33–35^ Because of this, the focus of care for children with HIV has shifted to treatment adherence, quality of life, and disease-associated morbidities such as cognitive impairment and psychiatric disorders.^33–36^

Multiple prior studies have demonstrated that cognitive impairment is common in children with HIV infection.^33–38^ With widespread use of ART, the major burden of cognitive impairment has shifted away from progressive encephalopathy toward milder forms of cognitive impairment.^35, 36, 39^ Of the numerous cognitive domains affected by HIV, executive function (EF) has been found to

be one of the most commonly affected domains in previous studies.^6, 40–43^ However, there is little data on executive function in children living with HIV in sub-Saharan Africa (where the majority of children living with HIV reside), and few published studies have evaluated whether executive function deficits in children living with HIV are clinically significant.^19, 35^

Prior studies in adults have demonstrated a relationship between poor executive function and poor ART adherence.^2, 44–50^ However, this relationship has been evaluated in only one pediatric study,^51^ which had inconclusive results. Taking ART as prescribed is a crucial skill for children and adolescents living with HIV to develop. ^27, 52, 53^ ART adherence is especially important for people living in sub-Saharan Africa because of limited availability of third-line and salvage ART regimens.^34, 35, 52, 54–56^ Adherence requires the recruitment of executive function because consistently taking medications involves developing and implementing a plan to adhere, remembering to adhere, trading immediate benefits for future gains, and developing strategies for adherence in the face of potential barriers.^2, 46, 47^ In younger children, parents or other relatives may supply some of these functions, but as children age into adolescence and adulthood, they are required to bear greater responsibility for adherence on their own.

The aim of the current study was to examine executive function among children and adolescents living with HIV compared to a control group of HIV-exposed uninfected (HEU) children and adolescents in Zambia. We hypothesized that both neuropsychological performance-based measures of executive function and parental ratings of executive function would be worse in participants living with HIV than in HEU controls. We further hypothesized that poor executive function measures at baseline would predict poor adherence over the subsequent one-year period.

## Patients and Methods

Methods for the HANDZ study, which involves various analyses including those in this paper, have previously been described ^57–61^ Briefly, children and adolescents living with HIV and a control group of HIV-exposed uninfected (HEU) children and adolescents were recruited in a prospective cohort study. Participants living with HIV were recruited from the Pediatric HIV/AIDS Center of Excellence (PCOE, a referral center for children living with HIV located in Lusaka, Zambia) during routine medication refill visits. Controls were recruited from the community by a trained community health worker. All participants were recruited from 2017-2018 and were 8-17 years old at time of enrollment. Participants were evaluated every three months after enrollment. We report results here from the baseline visit and the first year of follow-up.

### Inclusion/Exclusion Criteria

Children and adolescents living with HIV were excluded if they had not been receiving ART for at least one year prior to enrollment. Children and adolescents not living with HIV were included only if their mother had been infected with HIV during or prior to their pregnancy. Any potential participant with a history of central nervous system opportunistic infection or a chronic medical or psychiatric condition that would preclude participation in the study was excluded.

### Data Collected

Demographic and medical history information was collected using participant and parent interviews and chart review of electronic and paper medical records. Socioeconomic status (SES) was evaluated using an adapted version of the Multiple Indicator Cluster Survey-5 (MICS5, http://mics.unicef.org/tools). Measures of socioeconomic status were combined into a socioeconomic status index (SESI; see Mbewe et al., 2022 for details ^62^). Nutritional Status was assessed by transcribing participants’ growth curves from their medical records onto a standard WHO growth curve and then prospectively plotting growth at subsequent visits. Severe malnutrition, wasting, and stunting were defined according to standard WHO definitions.^63^

#### HIV History

For participants living with HIV, HIV history was obtained via participant and parent interview and through chart review. Measures of current and historical HIV disease severity (current and lowest recorded CD4 counts, current and worst recorded WHO stage, and number of previous hospitalizations) were combined into a disease severity index.

#### Adverse Life Events

A ten question survey developed in Zambia regarding adverse life events over the prior 12 months was administered to each participant. The survey includes questions related to illness and hospitalization of family members, change of residence, violence and abuse, and other negative life events. The survey generates an adverse life events score of 0-10, with higher scores indicating a greater number of adverse life events.

#### Executive Function

Full details of neuropsychological testing in the HANDZ study have previously been reported ^57^ Executive function was assessed through a combination of performance-based neuropsychological tests and participant and parent interviews. Parent interviews were used to complete the Behavior Rating Inventory of Executive Function, Second Edition, Parent Report (BRIEF2-PR) ^64^ The BRIEF is a standardized age-normed inventory for the assessment of executive function which has been widely utilized in studies in Zambia and elsewhere in sub-Saharan Africa ^7, 65^ Parent reports were utilized regardless of participant age. Age-adjusted T-scores for BRIEF subscales including the Emotional Regulation Index, Behavior Regulation Index, and Cognitive Regulation Index were computed, with higher scores indicating worse performance. A Z-score transformation of the BRIEF executive composite was used in logistic regression models for ease of interpretation. In addition, an executive function self-report measure developed by the authors for use in Africa, the Brief Executive Function Inventory (BEFI), was administered to participants.^57^ BEFI scores have a range of 0-10, with higher scores indicating worse performance. Performance-based neuropsychological testing is summarized in Table 1. Testing was conducted using a combination of standard measures and computerized testing using a custom NIH Toolbox Cognition Battery ^66–70^ Domain scores from all performance-based tests were combined into a global measure of executive function and transformed into a Z-score referred to as the NP Global Composite Score. Neuropsychological testing was conducted by PhD psychology students for standard measures and by a trained study nurse or research assistant for NIH Toolbox measures.

**Table 1:** Performance-based measures of executive function.

| **Domain** | **Test** |
| --- | --- |
| Set-shifting and cognitive flexibility | NIH Toolbox Directional Change Card Sort Test<sup>66, 69</sup><br>Wisconsin Card Sort Test 64 Item Version<br>DKEFS Trail Making Test Condition 4 |
| Attention/working memory | WISC5 Digit Span Total Score<br>California Verbal Learning Test Trial 1<br>NIH Toolbox List Sorting Working Memory Test<sup>66, 69</sup> |
| Response inhibition | Pencil Tapping Test<sup>71-73</sup><br>NIH Toolbox Flanker Test<sup>66, 69</sup> |

#### Adherence and Medication Supervision

Antiretroviral adherence was assessed at baseline and every 3 months using an established Pediatric HIV/AIDS Cohort Study (PHACS) instrument in addition to pill counts.^74^ Participants and their parents were asked to rate the participant’s adherence over the last 7 days and last 30 days. For the primary outcome variable, adherence was treated as a dichotomous variable (100% reported adherence vs. less than 100% reported adherence at any point over one year of follow-up). Viral load was assessed at baseline and after one year of follow-up. Participants with a viral load >50 copies/ml had repeat viral load testing scheduled for confirmation. Participants who were virally suppressed at baseline and had viral load >1000 copies/ml at one year were considered to have virologic treatment failure. All participants had standard adherence counseling, with escalation of adherence counseling in those participants with noted adherence issues or detectable viral load. Level of supervision was determined by asking participants to rate their level of supervision when taking ART on a three-point scale (from no supervision to complete supervision of all doses).

### Sample Size and Power

A sample size of 200 participants per group was planned based on simulation studies for each aim, with the goal of ensuring model stability and avoiding overfitting for regression models. Participants were overenrolled by 4% to account for potential loss to follow-up.

### Statistical Methods

Statistical analyses were performed using Stata 16 (Stata Corp LP, College Station, TX). Comparisons between groups were performed using chi-squared tests for categorical variables, T-tests for normally distributed continuous variables, and log rank tests for non-normally distributed continuous variables. Bivariable and multivariable linear regression models were constructed to evaluate determinants of executive function. Separate models were fit for participants with and without HIV. Logistic regression models were used to evaluate the relationship between executive function and adherence in participants with HIV. Causal models were pre-specified using Directed Acyclic Graphs (DAGs, see Figure 1). Multivariable models included variables prespecified for likely causal importance and indicated by the DAG as appropriate for adjustment. These variables included age, sex, and socioeconomic status index. Receiver operating characteristic (ROC) curves were constructed to investigate the performance characteristics of EF measures in predicting adherence problems (treated as a dichotomous variable) and virologic treatment failure over one year of follow-up for each level of parental supervision. Missing data were treated with pairwise deletion.

**Figure 1:**
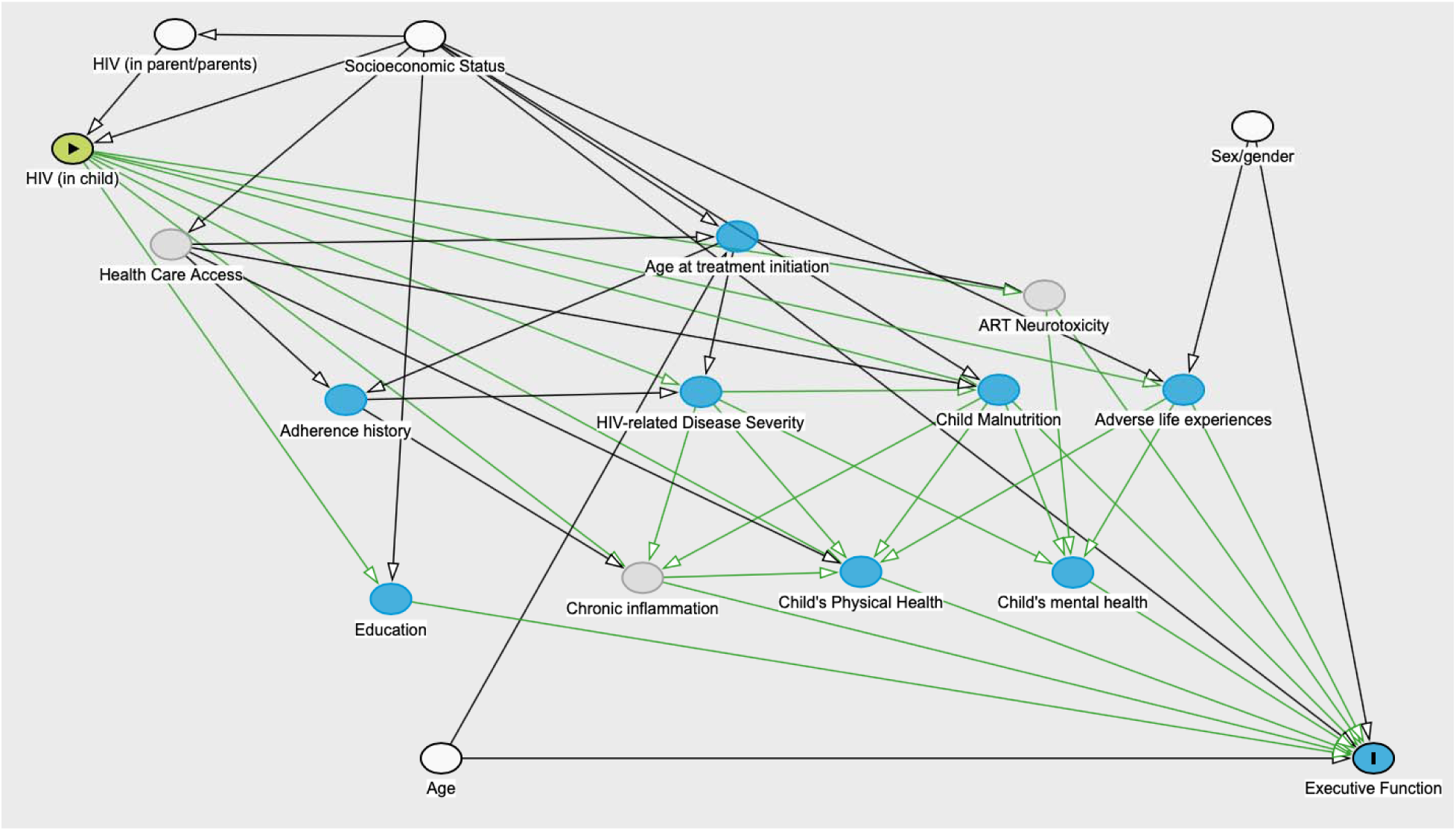
Directed Acyclic Graph (DAG) diagramming the causal model utilized in the study to estimate the effect of the exposure (HIV status) on the outcome of executive function. According to the DAG, the minimal sufficient adjustment set contains age, sex, parental HIV status, and socioeconomic status.

### Ethics Statement

This study was approved by the institutional review boards of the University of Rochester (protocol #00068985), the University of Zambia (reference #004-08-17), and the National Health Research Association of Zambia. Verbal and written informed consent was obtained from a parent for all participants for participation in the study, and verbal and written assent was obtained from all participants aged 12 or older.

## Results

### Demographics

A total of 416 participants were enrolled in the study. 29 participants were excluded from these analyses due to failure to complete neuropsychological testing, leaving 387 analyzable participants, including 205 participants living with HIV and 182 HEU controls. Participants living with HIV and HEU participants were demographically similar (see Table 2). However, participants living with HIV had slightly higher average socioeconomic status than controls, were more likely to attend school, and were less likely to report difficulty with food security. Additionally, participants with HIV were more likely to have a history of severe malnutrition. A detailed description of demographics and HIV-specific characteristics of this cohort and an explanation for the differences between groups has previously been published.^62^ All participants with HIV were treated with ART (median time on treatment: 7 years). Most participants with HIV had relatively high CD4 counts (median: 745, IQR: 537-975), and 83% had undetectable viral loads at baseline.

**Table 2:** Demographic/clinical characteristics of children living with HIV and controls. HEU: HIV-exposed Uninfected, TB: tuberculosis, IQR: interquartile range, WHO: World Health Organization. All data are presented as mean (SD) or n (%) unless otherwise specified. All p-values calculated using unpaired t-test or chi-squared test.

|  | Participants Living with HIV | HEU Participants | P-value |
| --- | --- | --- | --- |
| **Demographics:** |  |  |  |
| • Age at enrollment | 12 (2. 2) | 12 (2.7) | N/A |
| • Male sex (% male) | 114 (55%) | 87 (47%) | 0.15 |
| • Median socioeconomic status index (IQR) | 6 (5-8) | 5 (4-7) | <.001 |
| • Years of maternal education | 7.6 (3.4) | 6.4 (3.5) | <.001 |
| **Nutritional Status:** |  |  |  |
| • Low weight for age | 74 (36%) | 35 (20%) | <.001 |
| • Growth stunting | 66 (32%) | 32 (18%) | 0.001 |
| • History of severe malnutrition | 44 (21%) | 5 (2%) | 0.001 |
| **Child Physical Health:** |  |  |  |
| • Self-reported poor health | 21 (10%) | 11 (6%) | 0.135 |
| • History of hospitalization | 172 (83%) | 55 (27%) | <0.001 |
| • History of TB | 70 (33%) | 0 (0%) | <0.001 |
| **Education:** |  |  |  |
| • In school | 193 (92%) | 176 (85%) | .03 |
| • In private school | 100 (52%) | 57 (37%) | 0.01 |
| **HIV-specific Factors:** |  |  |  |
| • Current CD4 count | 772 (348) | N/A | N/A |
| • Nadir CD4 | 551 (347) | N/A | N/A |
| • History of CD4 count below 200 | 17 (8%) | N/A | N/A |
| • History of WHO stage 4 | 90 (43%) | N/A | N/A |
| • History of opportunistic infection other than TB | 16 (8%) | N/A | N/A |
| • Undetectable viral load at enrollment | 173 (83%) | N/A | N/A |
| • Age at ART initiation | <1 year: 23 (11%)<br>1-2 years: 33 (16%)<br>2-5 years: 70 (34%)<br>5-10 years: 56 (27%)<br>> 10 years: 26 (13%) | N/A | N/A |

### Executive Function in Participants Living with HIV Compared to HEU Controls and Comparison of Measures

Participants living with HIV performed significantly worse than HEU participants on all measures of executive function (see Table 3 and Figures 2a and 2b). These differences persisted after adjusting for age, sex, and socioeconomic status index (adjusted beta coefficient for BRIEF global executive composite: 2.6, 95% CI: 0.91 - 4.3, p = 0.003; adjusted beta coefficient for NP global composite: -0.52, 95% CI: -0.72 - -0.31, p < 0.001). Participants living with HIV also rated themselves as having poorer executive function on the BEFI (mean score: 2.3 vs. 1.7, p=0.004, adjusted beta coefficient: 0.66, 95% CI: 0.3 - 1.1, p = 0.002). Results from the BRIEF were mildly correlated with results from performance-based measures (Pearson correlation between BRIEF global executive composite and NP global composite (r) = -0.28, p<0.001).

**Figure 2:**
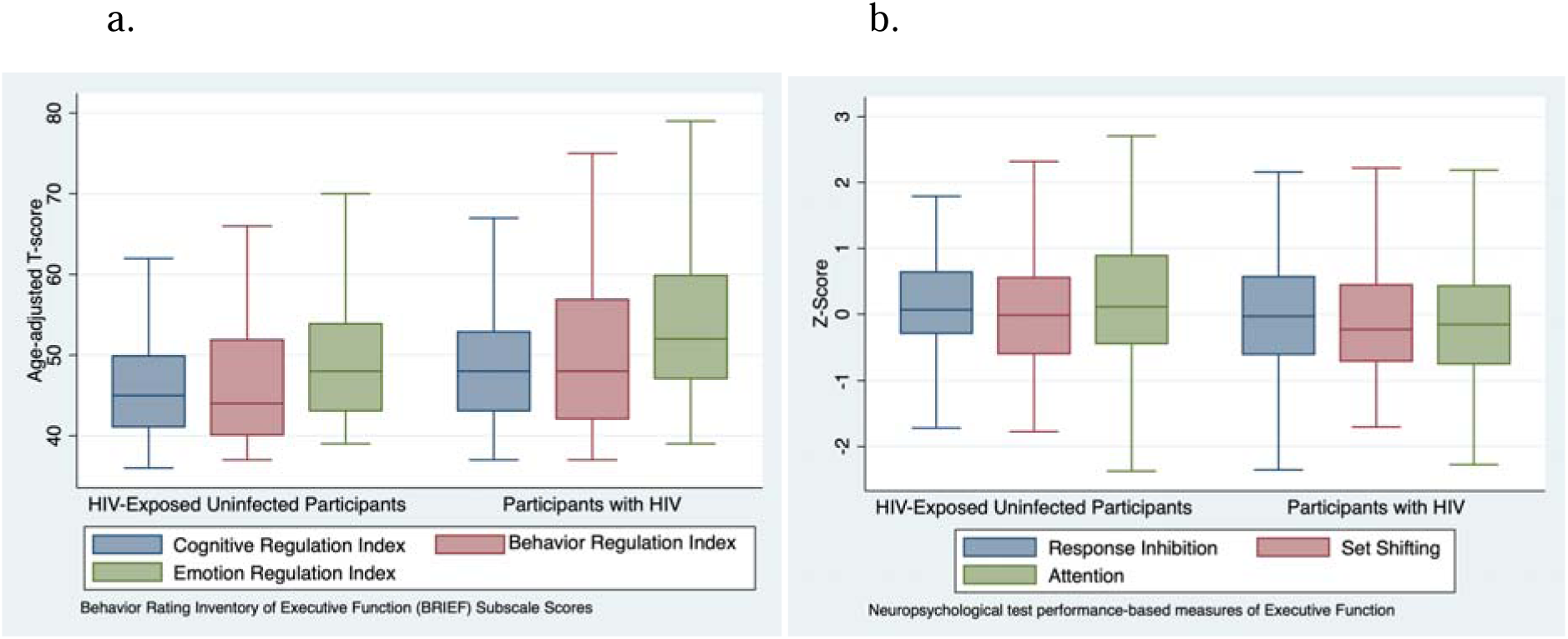
2a) BRIEF subscale scores by HIV status, with participants living with HIV demonstrating worse performance (higher scores) on all subscales. 2b) Performance-based test scores for multiple domains of executive function, with children living with HIV demonstrating worse performance (lower scores) than HIV-exposed uninfected controls.

**Table 3:** Executive function measures by HIV status.

| Measure | Participants Living with HIV | HIV-exposed Uninfected Participants | P-value |
| --- | --- | --- | --- |
| BRIEF measures (higher scores indicate worse performance) |  |  |  |
| Behavior regulation | 49.5 (48.2-50.8) | 46.7 (45.4-47.9) | 0.002 |
| Emotional regulation | 53.7 (52.4-54.9) | 49.8 (49.0-51.0) | <0.001 |
| Cognitive regulation | 48.9 (47.9-49.8) | 46.9 (45.8-48.1) | 0.01 |
| Global executive composite | 50.8 (49.5-51.6) | 47.8 (46.6-49.0) | <0.001 |
| Performance-based measures (higher scores indicate better performance) |  |  |  |
| Attention/working memory index | -0.17 (-0.30, 0.03) | 0.18 (0.04-0.32) | <0.001 |
| Set shifting | -0.17 (-0.30, -0.04) | 0.19 (0.04-0.34) | <0.001 |
| Response inhibition | -0.12 (-0.26-0.02) | 0.13 (0-0.27) | 0.01 |
| NP global composite | -0.20 (-0.34, -0.6) | 0.22 (0.08, 0.35) | <0.001 |

### Determinants of Executive Function in Participants Living with HIV

Separate bivariable and multivariable regression models were fit for the BRIEF global executive composite and the performance-based NP global composite scores. In the bivariable analyses for participants living with HIV, executive function was significantly associated with measures of nutritional status, socioeconomic status, and measures of HIV disease severity for both BRIEF and NP measures (see Table 4). Negative life events and younger age were associated with the BRIEF score but not with the NP global composite score. Sex was not associated with executive function on either measure.

**Table 4:**
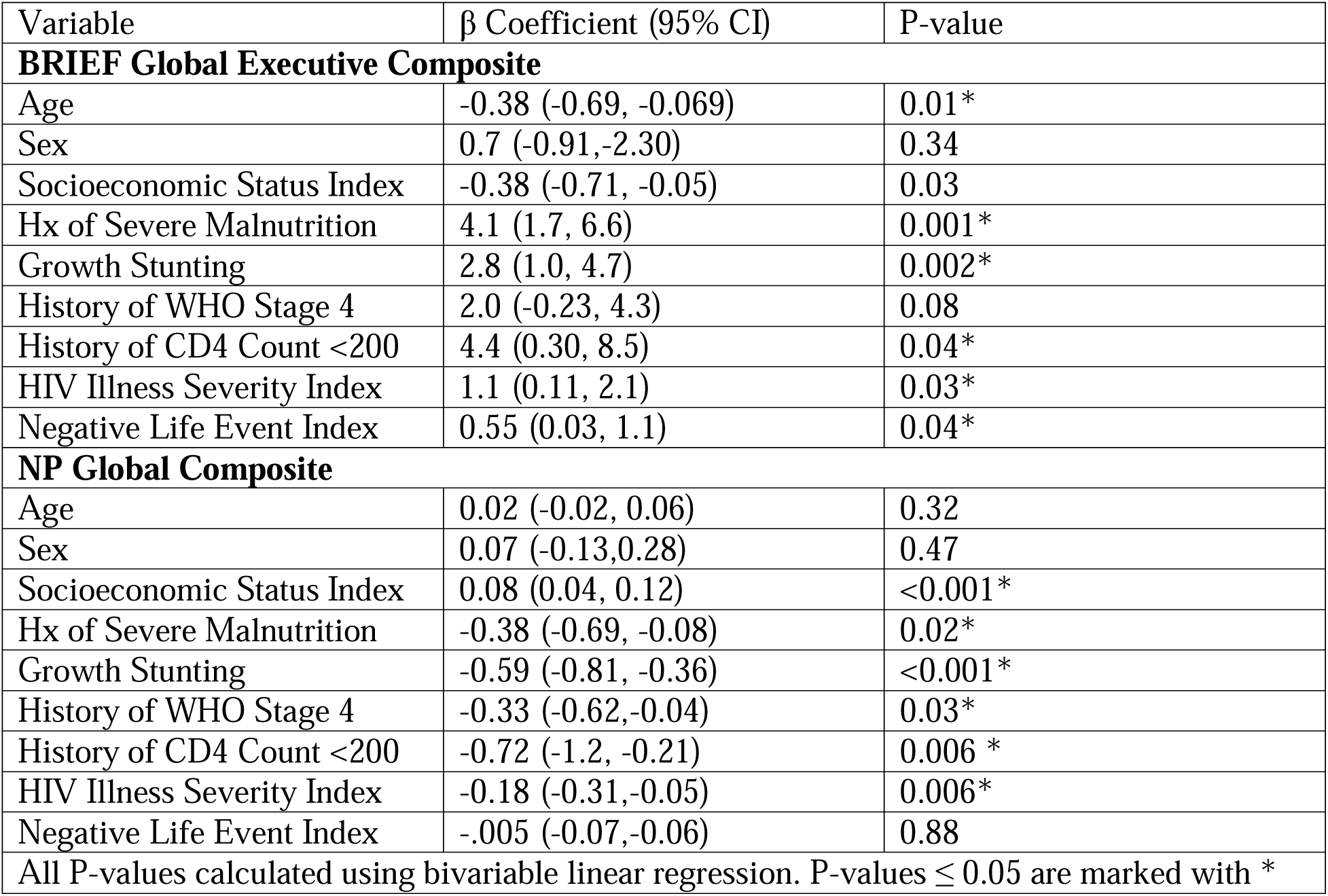
Determinants of executive function in participants living with HIV determined using bivariable linear regression.

| Variable | $\beta$ Coefficient (95% CI) | P-value |
| --- | --- | --- |
| **BRIEF Global Executive Composite** |  |  |
| Age | -0.38 (-0.69, -0.069) | 0.01* |
| Sex | 0.7 (-0.91,-2.30) | 0.34 |
| Socioeconomic Status Index | -0.38 (-0.71, -0.05) | 0.03 |
| Hx of Severe Malnutrition | 4.1 (1.7, 6.6) | 0.001* |
| Growth Stunting | 2.8 (1.0, 4.7) | 0.002* |
| History of WHO Stage 4 | 2.0 (-0.23, 4.3) | 0.08 |
| History of CD4 Count <200 | 4.4 (0.30, 8.5) | 0.04* |
| HIV Illness Severity Index | 1.1 (0.11, 2.1) | 0.03* |
| Negative Life Event Index | 0.55 (0.03, 1.1) | 0.04* |
| **NP Global Composite** |  |  |
| Age | 0.02 (-0.02, 0.06) | 0.32 |
| Sex | 0.07 (-0.13,0.28) | 0.47 |
| Socioeconomic Status Index | 0.08 (0.04, 0.12) | <0.001* |
| Hx of Severe Malnutrition | -0.38 (-0.69, -0.08) | 0.02* |
| Growth Stunting | -0.59 (-0.81, -0.36) | <0.001* |
| History of WHO Stage 4 | -0.33 (-0.62,-0.04) | 0.03* |
| History of CD4 Count <200 | -0.72 (-1.2, -0.21) | 0.006 * |
| HIV Illness Severity Index | -0.18 (-0.31,-0.05) | 0.006* |
| Negative Life Event Index | -.005 (-0.07,-0.06) | 0.88 |
| All P-values calculated using bivariable linear regression. P-values $\leq 0.05$ are marked with * | | |

For the multivariable models, all variables included in Table 4 were included. Age and sex were not significantly associated with outcomes in any of the final models. In the multivariable model for the BRIEF, SESI and adverse life event index had the strongest associations with EF (for SESI, adjusted β: -0.88, 95% CI: -1.1 - 0.05, p = 0.03; for adverse life event index, adjusted β: 1.2, 95% CI: 0.18 - 2.1, p= 0.02). In the multivariable model for the NP global composite score, growth stunting had the strongest association with EF (adjusted β: -0.38, 95% CI: -0.71 - 0.06, p = 0.02).

### Determinants of Executive Function in HEU Participants

For HEU participants, only history of severe malnutrition was significantly associated with the BRIEF global executive composite in the bivariable analysis (β = 10.1, 95% CI: 3.1 - 17.1, p = 0.005). This association remained significant in the multivariable model. For the NP global composite, growth stunting (β = -0.68, p < 0.001) and socioeconomic status (β = 0.11, p = 0.02) were the only variables with significant associations, and both remained significant in the multivariable model.

### Executive Function and Adherence

182 participants living with HIV had complete follow-up data at one year. 100% adherence was noted in 143 (79%) participants, while less than 100% adherence was noted in 39 out of the 182 participants (21%). There was no association between level of supervision and adherence. In a multivariable model controlling for SESI, age, and sex, the BRIEF executive composite at baseline predicted adherence problems over one year of follow-up (OR: 1.5, 95% CI: 1.02 - 2.2, p = 0.04). Performance-based and self-report measures of EF were not significantly associated with adherence problems.

In addition, worse executive function was associated with higher levels of parental supervision of adherence (see Figure 3). Participants reporting that parents observed all medication doses (compared to participants reporting at least some independence in taking medications) had higher scores on the BRIEF (mean BRIEF global executive composite: 53.0 vs. 50.0, p = 0.04) and BEFI (BEFI mean score: 2.8 vs. 2.2, p = 0.04) and lower scores on performance-based measures of EF (NP global composite: -0.62 vs. 0.08, p < 0.001). This relationship remained significant for the BRIEF cognitive regulation index and behavior regulation index but not the emotion regulation index and was significant for each individual performance-based measure of EF. ROC curve analysis suggested that all subscales of the BRIEF had moderate ability to predict poor adherence over the subsequent year of follow-up in participants with the lowest level of supervision (AUC 0.68 - 0.72, see Figure 4a). However, this was not the case in patients with moderate or greater levels of supervision (see Figure 4b). Executive was not significantly associated with virologic treatment failure for performance-based, parent-report, or self-report measures.

**Figure 3:**
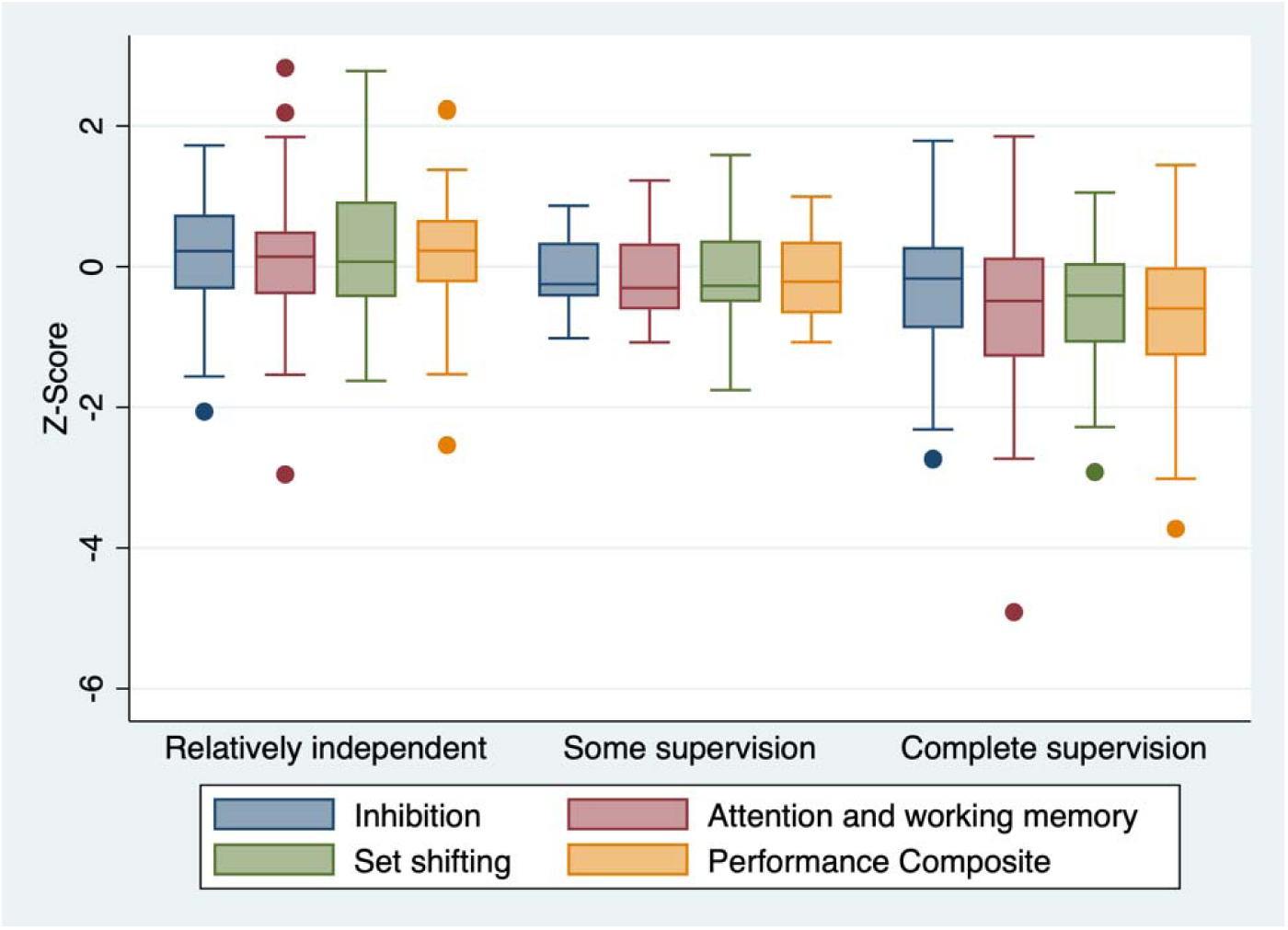
Performance-based measures of executive function by level of parental supervision of medication, demonstrating increasing levels of parental supervision in participants with poorer executive function.

**Figure 4:**
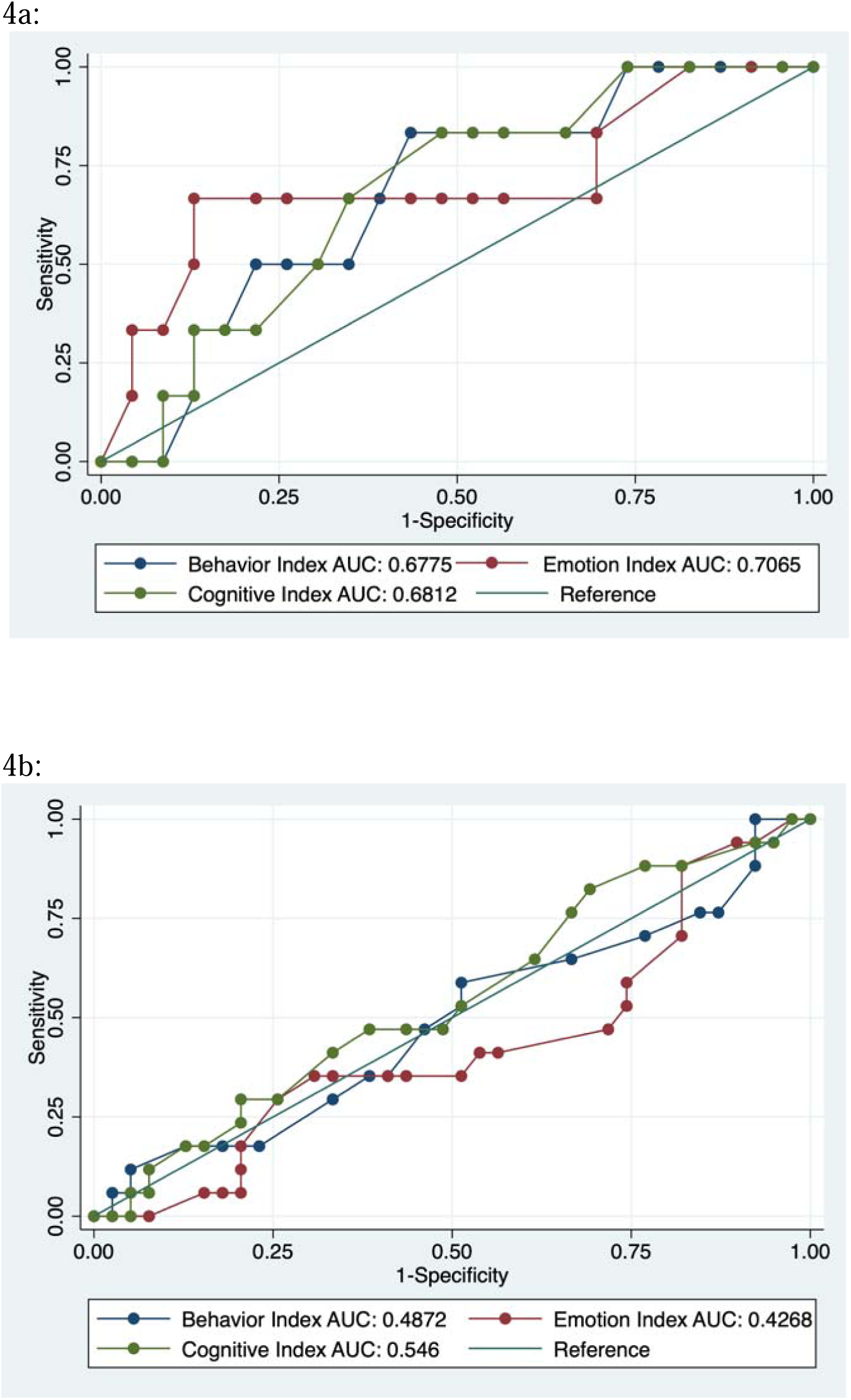
Receiver operator characteristic curves demonstrating the performance characteristics of BRIEF subscale scores in predicting adherence problems in participants with the lowest level of parental supervision (4a) and in those with complete parental supervision (4b). All subscales of the BRIEF had moderate ability (AUC: 0.68 to 0.71) to predict adherence problems in participants with minimal supervision but not in more highly supervised participants.

## Discussion

We examined the executive function of children living with HIV and not living with HIV as well as the effect of executive function on medication adherence among children with HIV in Lusaka, Zambia. Key findings included poorer performance on all measures of executive function in children living with HIV compared to HIV-exposed uninfected controls. The current study was not designed to investigate mechanisms of executive function impairment, but this EF impairment may be part of the spectrum of HIV-associated neurocognitive impairment. Interestingly, although measures of HIV disease severity were associated with EF, some of the strongest predictors of executive function were nutritional indicators, suggesting that some elements of EF impairment may be preventable with nutritional interventions and prevention of HIV-associated stunting and wasting. We identified a complex relationship between executive function, adherence, and level of parental supervision. Participants with worse executive function were more likely to have increased levels of parental supervision, suggesting that at least in a significant number of participants, poor executive function was compensated for by parents providing additional help with adherence. However, especially in participants with the lowest level of supervision, poor executive function assessed by parental report using the BRIEF predicted poor adherence over the following year. Future studies are necessary to determine whether problems with adherence become more of an issue as adolescents mature into adulthood and have to take additional responsibility for their own adherence, and we plan to address this as the HANDZ study continues.

In general, these findings are in line with most but not all prior studies of executive function in people living with HIV.^12–19^ Notably, a large study conducted in six African countries as part of the IMPAACT 1104 study^6–7^ noted deficits in performance-based measures of executive function, but not on parental ratings using the BRIEF. However, there was significant variability in results by site, with some sites reporting significant differences and others reporting no differences. Differences in our study compared to the 1104 study may be related to differences in the control group utilized or to the relative health of the participants living with HIV. While our cohort was relatively healthy, a substantially greater number of participants in our study started antiretroviral therapy relatively late and had a history of WHO stage 4 disease.

Results regarding determinants of EF in our cohort are generally comparable to results from prior studies, although we noted stronger relationships between EF and measures of HIV-specific disease severity than many prior studies did. The finding that socioeconomic status is strongly associated with EF is consistent with prior studies performed in Zambia^61,62^ and the United States.^3^ A relatively novel finding of our study is the strong relationship between malnutrition, stunting, and EF.

Our study contributes to findings from the PHACS study,^51^ which evaluated the relationship between HIV, executive function, and adherence in a U.S. cohort. Like the PHACS investigators, we noted that executive function deficits may be partly compensated for by increased levels of parental supervision.

### Limitations, Bias, and Generalizability

One notable limitation is the reliance on self-report and parental report measures of adherence, which have consistently been shown to underestimate rates of poor adherence. In the HANDZ cohort, there is a strong relationship between self-reported poor adherence and detectable viral load, suggesting that self-reported adherence is at least partially accurate, but objective measures of adherence would likely improve the accuracy of these assessments. In addition, given the relatively good rates of adherence noted in our study, we had inadequate power to implement more complex statistical techniques that could better illuminate the relationship between adherence and parental supervision. We plan to address these limitations in future longitudinal analyses after more non-adherence events occur. Additionally, our study was conducted at a single urban center in Zambia, and these findings may not generalize to other areas. In addition, we excluded participants with no available biological parents, and it is likely that the relationship between executive function and adherence is different in participants who are living with non-parent caregivers or in orphanages.

## Conclusion

Children and adolescents living with HIV had significantly poorer executive function compared to HIV-exposed uninfected controls. This is likely to be driven by a combination of factors, including social factors, health-related effects of HIV, and effects of HIV on the developing brain. The relationship between executive function and adherence is at least partially modified by parental supervision, as children with poor executive functioning are more likely to be highly supervised. However, especially in participants with the greatest level of independence, parental ratings of executive function were able to predict poor adherence over the subsequent year of follow-up. This finding raises concerns about the future as children age into adulthood and are required to take greater responsibility for medication adherence. Future longitudinal follow-up studies are planned as part of the HANDZ study to further investigate these issues.

## Contributors Statement

Sylvia Mwanza-Kabaghe: Dr. Mwanza-Kabaghe assisted in conceptualizing and designing the study, collected data, carried out data analysis, drafted and revised the manuscript, and approved the final manuscript as submitted. David R. Bearden: Dr. Bearden conceptualized and designed the study, collected data, carried out data analysis, assisted in all statistical analyses, assisted in drafting and revising the manuscript, and approved the final manuscript as submitted. All other authors assisted in designing and analyzing the study, assisted in drafting and revising the manuscript, and approved the final manuscript as submitted.

## Data Accessibility

Data is available upon reasonable request from David R. Bearden at

## Data Availability

All data produced in the present study are available upon reasonable request to the authors.

